# Safety and efficacy of a prehospital initiated protocol of nitrates plus non-invasive ventilation on prehospital and Emergency Department outcomes for acute cardiogenic pulmonary oedema

**DOI:** 10.1101/2021.10.17.21265081

**Authors:** Ian Howard, Nicholas Castle, Loua Al Shaikh, Robert Owen

**Affiliations:** Hamad Medical Corporation Ambulance Service, Hamad Medical Corporation, Doha, Qatar

## Abstract

**Background:** Acute heart failure is a common presentation to Emergency Departments (ED) the world over. Amongst the most common presenting signs and symptoms is dyspnoea due to acute pulmonary oedema, a life-threatening emergency that if left untreated or poorly managed. There is increasing evidence demonstrating improved outcomes following the use of vasodilators or non-invasive ventilation for these patients in the emergency setting. Consequently, the potential exists that initiating these therapies in the prehospital setting will similarly improve outcomes.

**Methods:** A historical cohort study was conducted to assess the effect of a prehospital initiated treatment protocol of nitrates plus non-invasive ventilation (NIV) versus regular therapy for severe cardiogenic APO on all-cause in-hospital mortality at 7 days, 30 days, and in total. Secondary outcomes included changes in EMS respiratory and haemodynamic parameters; admission status; length of stay; and emergency endotracheal intubation.

**Results:** The intervention led to an approximate 85% reduction in adjusted odds of mortality at 7 days compared to the regular therapy (AOR 0.15, 95% CI: 0.05 – 0.46, p = 0.001); approximate 80% reduction in odds of mortality at 30 days (AOR 0.19, 95% CI: 0.07 – 48, p < 0.0001); and Approximate 60% reduction in odds of total mortality (AOR 0.25, 95% CI: 0.12 – 0.56, p = 0.001).

**Conclusion:** The results of this analysis provide strong evidence of the potential synergistic benefits that can be achieved with the early implementation of a simple treatment protocol of prehospital administered nitrates and initiation of NIV for cardiogenic APO.

## INTRODUCTION

### Background

Acute heart failure (AHF) is a common presentation to Emergency Departments (ED) the world over, accounting for approximately one million ED attendances per year in the United States alone^1–3^. As many as 80% of these patients may potentially require admission to hospital, a burden that is likely to continue or worsen into the future with improved survival from cardiovascular disease and an ageing population^1–5^.

The heterogeneous manner in which patients in AHF present often makes their emergency management complex and problematic^5,6^. Amongst the most common presenting signs and symptoms is dyspnoea due to acute pulmonary oedema (APO), a life-threatening emergency that, if left untreated or poorly managed, may require tracheal intubation and mechanical ventilation, outcomes which are associated with a worse overall prognosis^5,7,8^. As many as 1 in 3 AHF patients will potentially present with APO, highlighting the importance of developing strategies aimed towards its early management^9^. Consequently, emergency care has a significant role to play in directly impacting mortality, morbidity, and hospital length of stay in this patient group.

### Importance

Historically, the acute management of APO consisted of supplemental oxygen therapy, diuretics and/or opiates^5,6,9^. However, the efficacy of these traditional therapies has been questioned in light of the scant and inconclusive results following their use^10–12^. There is increasing evidence demonstrating improved outcomes following the use of vasodilators^5,12^ or non-invasive ventilation (NIV)^5,13–15^ for these patients in the emergency setting. Traditionally, much of the literature’s emphasis has focused on delivering these therapies in the ED^5,6,9^. However, an accumulating body of evidence has highlighted the importance of “time to therapy” in managing cardiogenic APO^5,6,9^. Consequently, the emergency medical services (EMS) may have a potential role in reducing time to therapy and improving outcomes amongst this patient group.

With this in mind, this study aimed to investigate the effect of a prehospital initiated treatment protocol of nitrates and non-invasive ventilation on the prehospital and ED outcomes for patients presenting with acute pulmonary oedema of presumed cardiac origin.

## METHODS

### Study design and setting

The study was conducted within the Hamad Medical Corporation (HMC), the primary provider of secondary and tertiary healthcare in Qatar, and Hamad Medical Corporation Ambulance Service (HMCAS), a two-tiered national EMS provider with Ambulance Paramedic (AP) staffed ambulances and Critical Care Paramedic (CCP) staffed fast-response vehicles.

### Selection of Participants

The study utilized routinely collected clinical data extracted from the HMCAS and HMC electronic health record databases between December 2016 to August 2019. Patients were first identified in the HMCAS electronic patient care record (ePCR) database by “Provisional Diagnosis”. Due to diagnostic testing and assessment limitations in the prehospital setting, provisional diagnoses are primarily determined using either validated prehospital diagnostic criteria (e.g., ST elevation in myocardial infarction) or based on presenting signs and/or symptoms (e.g., bronchoconstriction in Asthma). Consequently, diagnosis of cardiogenic APO in the study service was primarily based on the presence of crackles/rales, elevated systolic blood pressure, and the absence of any other significant respiratory medical history (i.e.: asthma or COPD).

Provisional diagnoses in the HMCAS ePCR database utilize a diagnostic classification standard where diagnoses are categorized around similar diseases, disorders, injuries, and other related health conditions. For the purposes of this study, the EMS database was screened for the following provisional diagnoses:

- Cardiovascular: Acute Pulmonary Oedema
- Cardiovascular: Heart Failure

These cases were then linked with the ED database for cases with a primary, secondary, or tertiary diagnosis of:

- Acute pulmonary oedema
- Acute heart/cardiac failure
- Congestive heart/cardiac failure

From this target population, the following criteria were used for inclusion into the study:

#### Inclusion criteria

- Patients >= 18 years old
- Patients who have had at least two blood pressure recordings, a reading at patient arrival of EMS, and a reading at patient ED handover
- Patients treated and transported by HMCAS between Dec 2016 – Aug 2019

#### Exclusion criteria

- Patients who refused transport
- Patient with an out of hospital cardiac arrest
- Patients transferred between health facilities
- Non-cardiogenic acute pulmonary oedema (e.g.: drowning)

### Interventions

The study employed a retrospective cohort design that used both a historical and concurrent control group. Following an internal audit of the prehospital management of APO and review of the current scientific literature, an updated Clinical Practice Guideline (CPG) was developed for the treatment and transportation of cardiogenic APO patients by EMS (Figure 1). Prior to implementation of the CPG, the historical approach had allowed for discretion on the part of the treating clinician in the management of these patients. With the introduction of the CPG, clinicians were mandated to provide at the minimum, a treatment protocol consisting of early high dose sublingual Glyceryl Trinitrate (GTN) in combination with the initiation of Continuous Positive Airway Pressure (CPAP) NIV. The protocol was limited to the CCP scope of practice, and consequently, only patients who had a CCP arrive at patient side had the potential to receive the treatment protocol. For the purposes of this study, patients who received the treatment protocol made up the intervention group, and those who received the historical, “regular therapy” were classified as the reference/control group. Patients who were not attended to by a CCP, and consequently received no active therapy were included as an additional concurrent control group.

**Figure 1:**
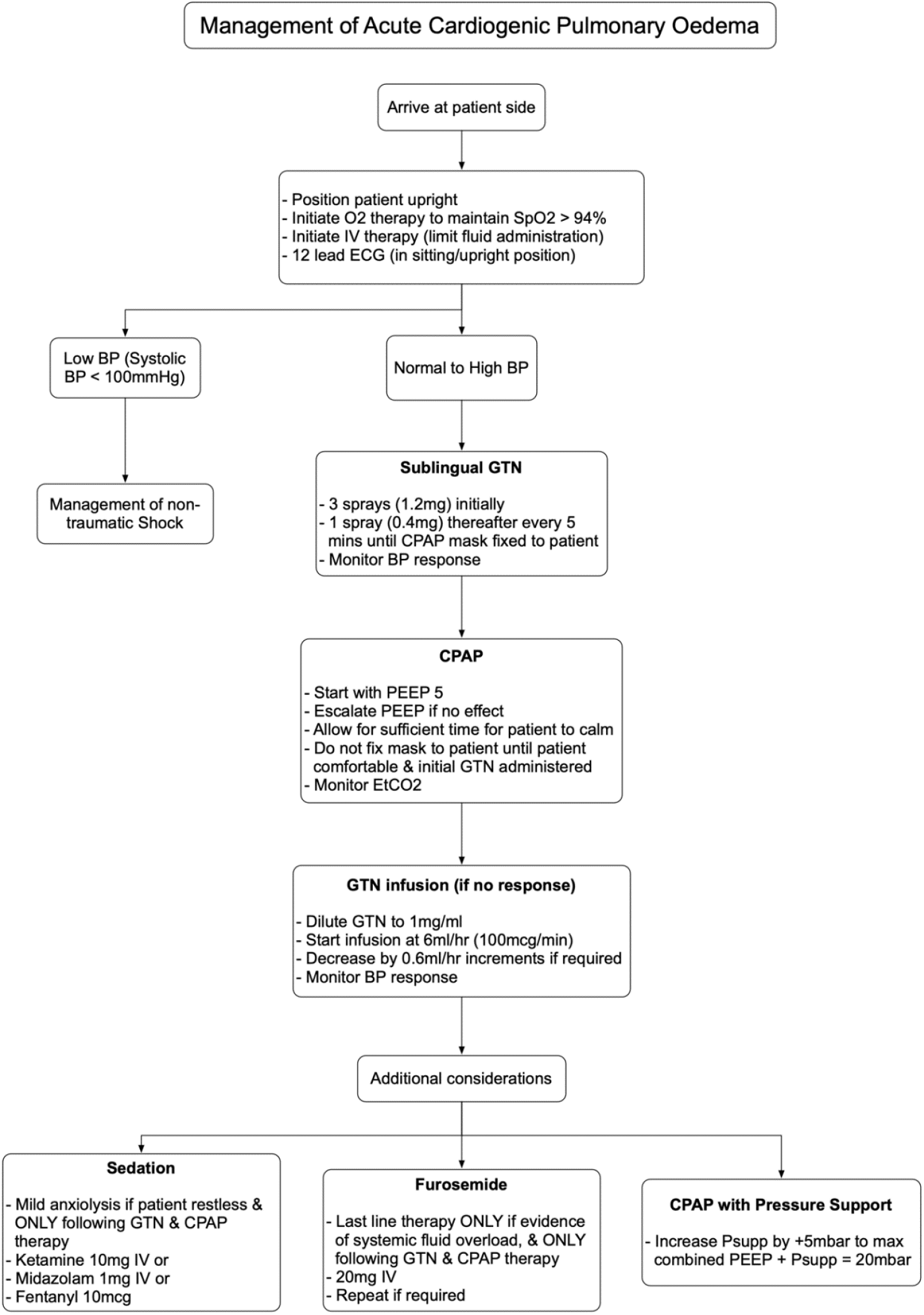
Clinical Practice.

### Measurements and Outcomes

Data collection and analysis were split into two parts. Part One focused on the extraction and analysis of EMS data and included EMS call-time intervals, EMS patient demographics; EMS vital signs; EMS interventions performed, and medications administered. Part Two focused on the linkage of EMS data with the relevant hospital data for each specific ED encounter and admission. The data collected included: ED interventions performed, medications administered, ED patient disposition, ED length of stay, ED diagnosis, hospital length of stay and hospital mortality. The primary outcomes under study were all-cause in-hospital mortality at 7-days, 30-days, and total in-hospital mortality. Secondary outcomes included changes in EMS respiratory and haemodynamic parameters; admission status; ED and in-patient length of stay; and a composite measure of non-cardiac arrest emergency endotracheal intubation (ETI) i.e.: emergency intubation performed in either the prehospital environment or the ED not in the context of cardiac arrest.

All EMS vital signs were captured via a combined ECG/vital signs monitor and automatically transmitted to the paired transporting vehicle’s ePCR. Similarly, call times and intervals were captured electronically using a computer-aided dispatch system and uploaded automatically without interference. EMS medications and procedures are mandated fields in the ePCR that utilize prompts to ensure a minimum level of information is captured regarding each medication or procedure. Overall, data for both intervention and exposure groups were collected from equal sources.

### Analysis

Measures of association between outcome and intervention status; and between multiple categorical exposure variables and intervention status were analysed using the chi-square test for association and reported using frequencies, percentages and 95% CIs. Comparison of changes in haemodynamic parameters between control and intervention groups with continuous data were assessed and compared using one-way ANOVA and reported using the variable mean, standard deviation (SD) and 95% CI.

Multivariate logistic regression was used to assess for odds of the primary and secondary outcomes by intervention status in both the prehospital and ED setting, adjusting for multiple variables of interest. A forward modelling strategy was employed for all multivariable analysis, starting with a base model of the intervention of interest. The study group of patients who received no therapy were used as the primary control group. An adjusted model with all the potential confounders considered *a priori* was then analysed and compared to the base model. A final model was produced comparing the intervention/protocol group with the regular therapy group as a reference, again adjusted for *a priori* confounders.

For the purpose of this study, age, gender, and a starting systolic blood pressure greater than 140 mmHg were considered as *a priori* confounding variables. The blood pressure component was included given it’s use, both historically and in the current CPG, as basic criteria for initiating therapy in this patient cohort. A >10% change in ORs was deemed to be meaningful confounding, and a doubling of confidence intervals was considered strong evidence of collinearity. Outputs for all models were reported using adjusted ORs and their 95% CI. All statistical analyses were performed using Stata version 15.1 (Stata Corporation, College Station, TX, USA). Ethical approval was obtained from the HMC Medical Research Committee and HMCAS Group Research Oversight Committee (Ref: MRC-01-18-113).

## RESULTS

### EMS analysis

A total of 1749 patients met inclusion criteria into the study, with all cases included in the EMS analysis, divided between the “No therapy” group (n = 680), “Regular therapy” group (n = 480) and “Protocol” group (n = 589). Patients were typically male and aged between 60–69 (Table 1). The differences in EMS time intervals were generally marginal between groups. Most notably, mean scene times were increased by approximately two minutes in the Protocol group compared to the other groups, likely as a consequence of the delivery of the treatment protocol (0:31:11 (0:13:47) vs 0:31:55 (0:12:49) vs 0:33:38 (0:13:22), p = 0.01). However, this is unlikely to have had any negative clinical consequence given the small difference and generally equal standard deviation in mean Scene times across the groups. Similarly, Patient Contact Times were generally equitable across the groups, with the differences in Total Case Time likely due to variations in response and transport times

**Table 1:**
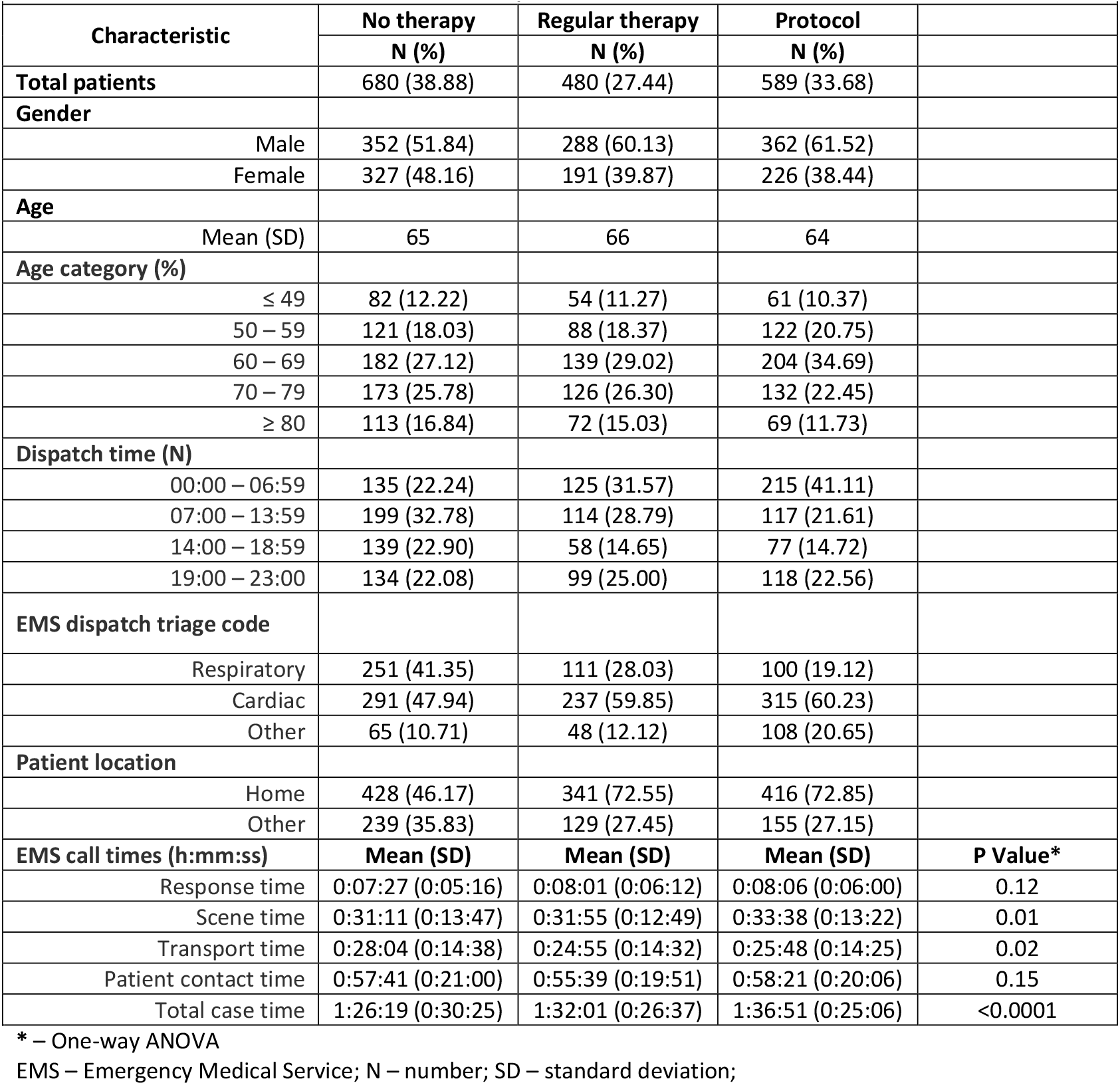
EMS patient characteristics.

| Table 1: EMS patient characteristics |  |  |  |  |
| --- | --- | --- | --- | --- |
| Characteristic | No therapy | Regular therapy | Protocol |  |
|  | N (%) | N (%) | N (%) |  |
| Total patients | 680 (38.88) | 480 (27.44) | 589 (33.68) |  |
| Gender |  |  |  |  |
| Male | 352 (51.84) | 288 (60.13) | 362 (61.52) |  |
| Female | 327 (48.16) | 191 (39.87) | 226 (38.44) |  |
| Age |  |  |  |  |
| Mean (SD) | 65 | 66 | 64 |  |
| Age category (%) |  |  |  |  |
| ≤ 49 | 82 (12.22) | 54 (11.27) | 61 (10.37) |  |
| 50 – 59 | 121 (18.03) | 88 (18.37) | 122 (20.75) |  |
| 60 – 69 | 182 (27.12) | 139 (29.02) | 204 (34.69) |  |
| 70 – 79 | 173 (25.78) | 126 (26.30) | 132 (22.45) |  |
| ≥ 80 | 113 (16.84) | 72 (15.03) | 69 (11.73) |  |
| Dispatch time (N) |  |  |  |  |
| 00:00 – 06:59 | 135 (22.24) | 125 (31.57) | 215 (41.11) |  |
| 07:00 – 13:59 | 199 (32.78) | 114 (28.79) | 117 (21.61) |  |
| 14:00 – 18:59 | 139 (22.90) | 58 (14.65) | 77 (14.72) |  |
| 19:00 – 23:00 | 134 (22.08) | 99 (25.00) | 118 (22.56) |  |
| EMS dispatch triage code |  |  |  |  |
| Respiratory | 251 (41.35) | 111 (28.03) | 100 (19.12) |  |
| Cardiac | 291 (47.94) | 237 (59.85) | 315 (60.23) |  |
| Other | 65 (10.71) | 48 (12.12) | 108 (20.65) |  |
| Patient location |  |  |  |  |
| Home | 428 (46.17) | 341 (72.55) | 416 (72.85) |  |
| Other | 239 (35.83) | 129 (27.45) | 155 (27.15) |  |
| EMS call times (h:mm:ss) | Mean (SD) | Mean (SD) | Mean (SD) | P Value* |
| Response time | 0:07:27 (0:05:16) | 0:08:01 (0:06:12) | 0:08:06 (0:06:00) | 0.12 |
| Scene time | 0:31:11 (0:13:47) | 0:31:55 (0:12:49) | 0:33:38 (0:13:22) | 0.01 |
| Transport time | 0:28:04 (0:14:38) | 0:24:55 (0:14:32) | 0:25:48 (0:14:25) | 0.02 |
| Patient contact time | 0:57:41 (0:21:00) | 0:55:39 (0:19:51) | 0:58:21 (0:20:06) | 0.15 |
| Total case time | 1:26:19 (0:30:25) | 1:32:01 (0:26:37) | 1:36:51 (0:25:06) | <0.0001 |

\* – One-way ANOVA
EMS – Emergency Medical Service; N – number; SD – standard deviation;

There were several differences in initial presentation between groups regarding primary EMS vital signs assessment (Table 2). Patients in the Protocol group generally had a higher initial heart rate, systolic blood pressure and lower SpO_2_ compared to the No Therapy group yet were relatively well matched with the Regular Therapy group across haemodynamic parameters. This indicates a general increased disease severity in the Regular therapy and Protocol groups, and further reinforces the need for intervention, compared with the No therapy group. In light disease severity, there was a stronger tendency towards recovery of vital signs to normal limits amongst the Protocol group compared with the regular therapy group, where greater reductions in high blood pressures and heart rates, and greater increases in oxygen saturation were observed.

**Table 2:**
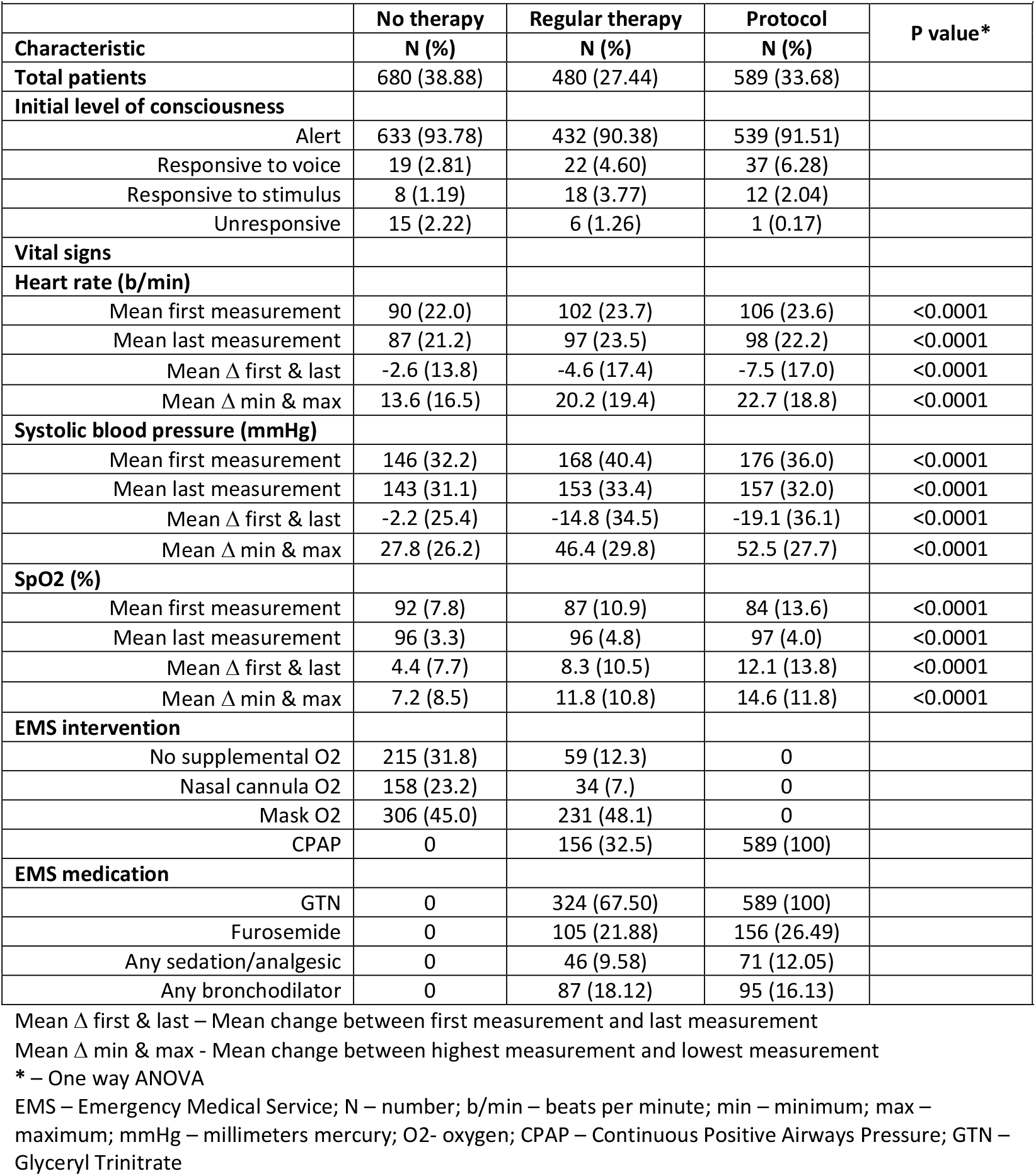
Initial EMS patient presentation and management.

EMS – Emergency Medical Service; N – number; SD – standard deviation;
| Table 2: Initial EMS patient presentation and management |  |  |  |  |
| --- | --- | --- | --- | --- |
|  | No therapy | Regular therapy | Protocol | P value* |
| Characteristic | N (%) | N (%) | N (%) |  |
| Total patients | 680 (38.88) | 480 (27.44) | 589 (33.68) |  |
| Initial level of consciousness |  |  |  |  |
| Alert | 633 (93.78) | 432 (90.38) | 539 (91.51) |  |
| Responsive to voice | 19 (2.81) | 22 (4.60) | 37 (6.28) |  |
| Responsive to stimulus | 8 (1.19) | 18 (3.77) | 12 (2.04) |  |
| Unresponsive | 15 (2.22) | 6 (1.26) | 1 (0.17) |  |
| Vital signs |  |  |  |  |
| Heart rate (b/min) |  |  |  |  |
| Mean first measurement | 90 (22.0) | 102 (23.7) | 106 (23.6) | <0.0001 |
| Mean last measurement | 87 (21.2) | 97 (23.5) | 98 (22.2) | <0.0001 |
| Mean $\Delta$ first & last | -2.6 (13.8) | -4.6 (17.4) | -7.5 (17.0) | <0.0001 |
| Mean $\Delta$ min & max | 13.6 (16.5) | 20.2 (19.4) | 22.7 (18.8) | <0.0001 |
| Systolic blood pressure (mmHg) |  |  |  |  |
| Mean first measurement | 146 (32.2) | 168 (40.4) | 176 (36.0) | <0.0001 |
| Mean last measurement | 143 (31.1) | 153 (33.4) | 157 (32.0) | <0.0001 |
| Mean $\Delta$ first & last | -2.2 (25.4) | -14.8 (34.5) | -19.1 (36.1) | <0.0001 |
| Mean $\Delta$ min & max | 27.8 (26.2) | 46.4 (29.8) | 52.5 (27.7) | <0.0001 |
| SpO2 (%) |  |  |  |  |
| Mean first measurement | 92 (7.8) | 87 (10.9) | 84 (13.6) | <0.0001 |
| Mean last measurement | 96 (3.3) | 96 (4.8) | 97 (4.0) | <0.0001 |
| Mean $\Delta$ first & last | 4.4 (7.7) | 8.3 (10.5) | 12.1 (13.8) | <0.0001 |
| Mean $\Delta$ min & max | 7.2 (8.5) | 11.8 (10.8) | 14.6 (11.8) | <0.0001 |
| EMS intervention |  |  |  |  |
| No supplemental O2 | 215 (31.8) | 59 (12.3) | 0 |  |
| Nasal cannula O2 | 158 (23.2) | 34 (7.) | 0 |  |
| Mask O2 | 306 (45.0) | 231 (48.1) | 0 |  |
| CPAP | 0 | 156 (32.5) | 589 (100) |  |
| EMS medication |  |  |  |  |
| GTN | 0 | 324 (67.50) | 589 (100) |  |
| Furosemide | 0 | 105 (21.88) | 156 (26.49) |  |
| Any sedation/analgesic | 0 | 46 (9.58) | 71 (12.05) |  |
| Any bronchodilator | 0 | 87 (18.12) | 95 (16.13) |  |

Mean $\Delta$ first & last – Mean change between first measurement and last measurement
Mean $\Delta$ min & max - Mean change between highest measurement and lowest measurement
\* – One way ANOVA
EMS – Emergency Medical Service; N – number; b/min – beats per minute; min – minimum; max – maximum; mmHg – millimeters mercury; O2- oxygen; CPAP – Continuous Positive Airways Pressure; GTN – Glyceryl Trinitrate

Evidence of the historical approach to the treatment of this patient group is apparent in the delivery of prehospital interventions, where approximately a third of patients in the Regular therapy group received CPAP, and approximately two thirds received a dose of GTN, compared to the Protocol group where these became minimum mandated interventions for all patients following the introduction of the new CPG. A likely consequence of the greater expected use of CPAP was the administration of a higher proportion of some form of anxiolysis in the Protocol group, compared with the Regular therapy group (Regular therapy – 9.58% vs Protocol – 12.05%). It is therefore arguable that this could have contributed to the greater changes in heart rates and blood pressures seen in the Protocol group compared to the other groups.

### ED analysis

In terms of hospital outcomes, a higher proportion of the Protocol group were discharged home directly from the ED (35.73%), compared with the regular therapy group (34.78%) (Table 3). Despite this, however, the Protocol group required a marginally longer mean stay in the ED (1.68 days), compared with the Regular therapy group (1.44 days). From an inpatient perspective, while a higher proportion of the Protocol group required general hospital admission (58.24%), compared with the regular therapy group (55.94%), fewer patients in the Protocol group required admission to ICU (13.92%) compared to the Regular therapy group (17.97). Overall, the Protocol group had the lowest mean length of hospital stay out of all three groups (8.88 days, vs No therapy – 12.96 days, vs Regular therapy – 13.08 days).

**Table 3:** ED/Hospital patient characteristics – distribution and crude association with intervention status.

|  |  |  |  |  |
| --- | --- | --- | --- | --- |
| **Characteristic** | **No therapy**<br>**N (%)** | **Regular therapy**<br>**N (%)** | **Protocol**<br>**N (%)** | **P value\*** |
| **Total patients** | 517 | 345 | 431 |  |
| **Patient disposition** |  |  |  |  |
| Discharged home from ED | 199 (38.49) | 120 (34.78) | 154 (35.73) | 0.01 |
| Absconded/DAMA/LAMA | 40 (7.74) | 28 (8.12) | 28 (6.50) |  |
| Discharge/transfer to other facility | 7 (1.35) | 6 (1.74) | 7 (1.62) |  |
| Discharged home from ward | 238 (46.03) | 164 (47.54) | 233 (54.06) |  |
| Died | 33 (6.38) | 27 (7.83) | 9 (2.09) |  |
| **In-hospital admission** | 275 (53.19) | 193 (55.94) | 251 (58.24) | 0.29 |
| **Admitted to ICU** | 56 (10.83) | 62 (17.97) | 60 (13.92) | 0.01 |
| **Mean length of stay (days)** | **Mean (SD)** | **Mean (SD)** | **Mean (SD)** | **P value\*\*** |
| Emergency department | 1.16 (1.64) | 1.44 (1.86) | 1.68 (1.85) | <0.0001 |
| Inpatient | 11.80 (78.38) | 11.63 (47.18) | 7.20 (22.23) | 0.399 |
| Total | 12.96 (78.52) | 13.08 (47.18) | 8.88 (22.23) | 0.467 |

\* - Chi2 test for association
\*\* – One way ANOVA
ED – Emergency department; N – number; DAMA – discharged against medical advice; LAMA – Left against medical advice; ICU – Intensive care unit; SD – Standard deviation

### Multivariable analysis

While there were fewer emergency intubations (EMS/ED composite) in the Protocol group (3.94%), compared with the No therapy group (4.26%) and Regular therapy group (4.93%), there was little evidence to suggest differences in either crude or adjusted odds of emergency intubation in the Protocol group compared with either both treatment groups (AOR 0.57, 95% CI: 0.26 – 1.22, p = 0.14) or between the Regular therapy group and Protocol group (AOR 0.79, 95% CI: 0.38 – 1.66, p = 0.53) (Table 4).

**Table 4:**
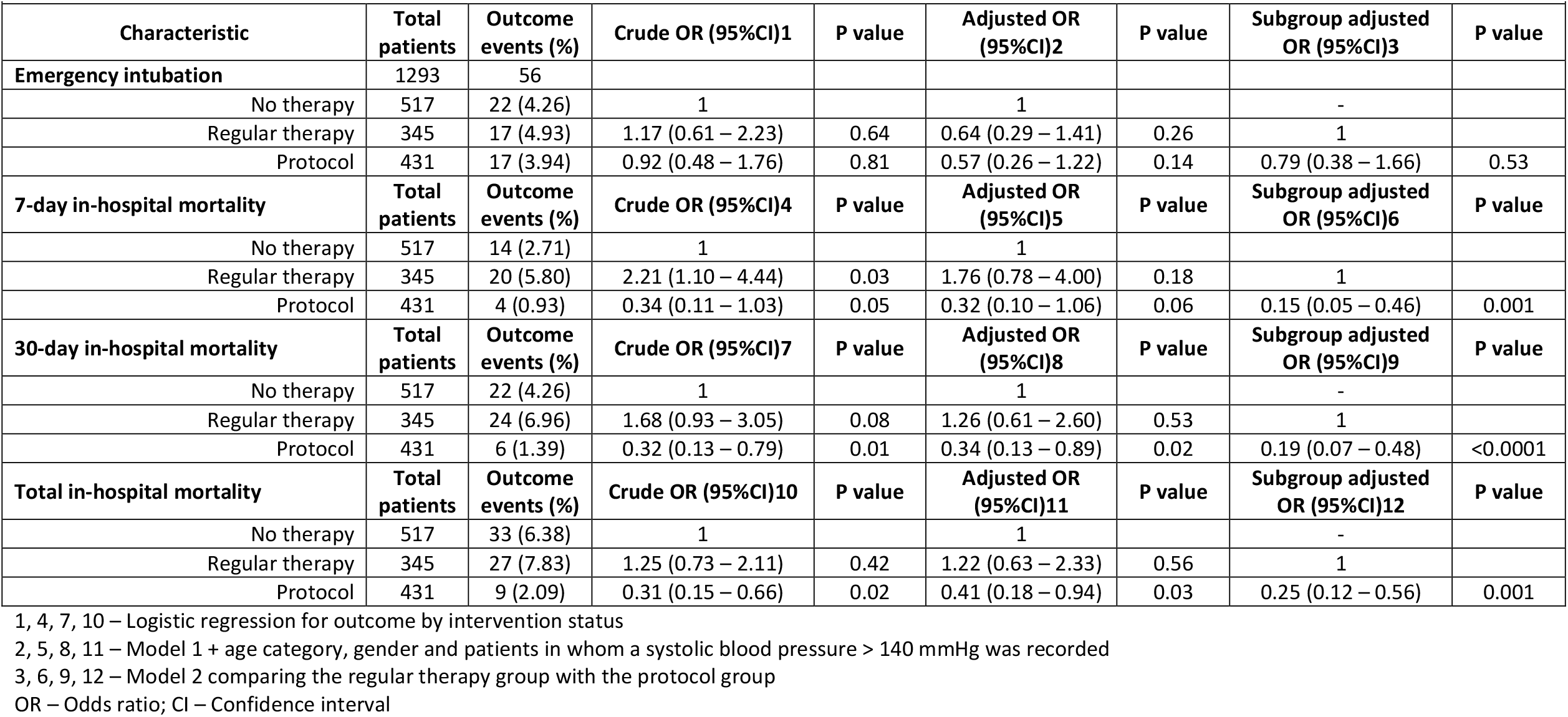
Multivariable analysis for the effect of the treatment protocol on primary and secondary outcomes.

|  |  |  |  |  |  |  |  |  |
| --- | --- | --- | --- | --- | --- | --- | --- | --- |
| Characteristic | Total patients | Outcome events (%) | Crude OR (95%CI)1 | P value | Adjusted OR (95%CI)2 | P value | Subgroup adjusted OR (95%CI)3 | P value |
| **Emergency intubation** | 1293 | 56 |  |  |  |  |  |  |
| No therapy | 517 | 22 (4.26) | 1 |  | 1 |  | - |  |
| Regular therapy | 345 | 17 (4.93) | 1.17 (0.61 – 2.23) | 0.64 | 0.64 (0.29 – 1.41) | 0.26 | 1 |  |
| Protocol | 431 | 17 (3.94) | 0.92 (0.48 – 1.76) | 0.81 | 0.57 (0.26 – 1.22) | 0.14 | 0.79 (0.38 – 1.66) | 0.53 |
| **7-day in-hospital mortality** | **Total patients** | **Outcome events (%)** | **Crude OR (95%CI)4** | **P value** | **Adjusted OR (95%CI)5** | **P value** | **Subgroup adjusted OR (95%CI)6** | **P value** |
| No therapy | 517 | 14 (2.71) | 1 |  | 1 |  |  |  |
| Regular therapy | 345 | 20 (5.80) | 2.21 (1.10 – 4.44) | 0.03 | 1.76 (0.78 – 4.00) | 0.18 | 1 |  |
| Protocol | 431 | 4 (0.93) | 0.34 (0.11 – 1.03) | 0.05 | 0.32 (0.10 – 1.06) | 0.06 | 0.15 (0.05 – 0.46) | 0.001 |
| **30-day in-hospital mortality** | **Total patients** | **Outcome events (%)** | **Crude OR (95%CI)7** | **P value** | **Adjusted OR (95%CI)8** | **P value** | **Subgroup adjusted OR (95%CI)9** | **P value** |
| No therapy | 517 | 22 (4.26) | 1 |  | 1 |  | - |  |
| Regular therapy | 345 | 24 (6.96) | 1.68 (0.93 – 3.05) | 0.08 | 1.26 (0.61 – 2.60) | 0.53 | 1 |  |
| Protocol | 431 | 6 (1.39) | 0.32 (0.13 – 0.79) | 0.01 | 0.34 (0.13 – 0.89) | 0.02 | 0.19 (0.07 – 0.48) | <0.0001 |
| **Total in-hospital mortality** | **Total patients** | **Outcome events (%)** | **Crude OR (95%CI)10** | **P value** | **Adjusted OR (95%CI)11** | **P value** | **Subgroup adjusted OR (95%CI)12** | **P value** |
| No therapy | 517 | 33 (6.38) | 1 |  | 1 |  | - |  |
| Regular therapy | 345 | 27 (7.83) | 1.25 (0.73 – 2.11) | 0.42 | 1.22 (0.63 – 2.33) | 0.56 | 1 |  |
| Protocol | 431 | 9 (2.09) | 0.31 (0.15 – 0.66) | 0.02 | 0.41 (0.18 – 0.94) | 0.03 | 0.25 (0.12 – 0.56) | 0.001 |

1, 4, 7, 10 – Logistic regression for outcome by intervention status
2, 5, 8, 11 – Model 1 + age category, gender and patients in whom a systolic blood pressure > 140 mmHg was recorded
3, 6, 9, 12 – Model 2 comparing the regular therapy group with the protocol group
OR – Odds ratio; CI – Confidence interval

From an all cause in-hospital mortality perspective, there were fewer events in the Protocol group at 7 days (0.93%, vs No therapy – 2.71%, vs Regular therapy – 5.80%); 30 days (1.39%, vs No therapy – 4.26%, vs Regular therapy – 4.26%), and in total (2.09%, vs No therapy – 6.38%, vs Regular therapy – 7.83%) compared to the other two groups. This translated to an approximate 68% reduced adjusted odds at 7 days, compared with both groups (AOR 0.32, 95% CI: 0.10 – 1.06, p = 0.06) and an approximate 85% reduction in adjusted odds compared to the regular therapy group alone (AOR 0.15, 95% CI: 0.05 – 0.46, p = 0.001). This reduced adjusted odds was sustained at both 30 days compared with both groups (AOR 0.34, 95% CI: 0.13 – 0.89, p = 0.02), or the Regular therapy group alone (AOR 0.19, 95% CI: 0.07 – 48, p < 0.0001); and in total, again compared with both reference groups (AOR 0.41, 95% CI: 0.18 – 0.94, p = 0.03) or the Regular therapy alone (AOR 0.25, 95% CI: 0.12 – 0.56, p = 0.001).

## LIMITATIONS

Inclusion in the study was primarily based on EMS provisional diagnosis. This was purposely carried out to measure the impact of the treatment protocol from an EMS perspective. Consequently, the potential exists that a proportion of patients were incorrectly included into and/or rejected from the study. In order to account for this, the EMS diagnosis was matched with ED diagnostic criteria to identify patients with APO of primarily assumed cardiovascular origin.

Data regarding the presence of underlying comorbidities was not available, and consequently their contribution towards the study outcomes was not assessed or controlled for. Similarly, data regarding lifestyle risk factors such as smoking, or obesity were not available, and their subsequent impact was not assessed. It is acknowledged that these risk factors could have potentially impacted the study outcomes and therefore remain important avenues for future research. Towards this, as highlighted above, diagnosis of APO in the study service was primarily based on the presence of crackles/rales, elevated systolic blood pressure, and the absence of any other significant respiratory medical history (i.e.: asthma or COPD). Therefore, it is assumed that the patients included in this study were generally of Killip class 2 or 3.

There were proportions of patients for which ED/Hospital data could not be retrieved or could not be linked to the EMS data set. The potential exists that the hospital data for these patients could have differed through either their exposure, intervention, or outcome status. However, further investigation into reasons for this missingness found them to be primarily related to incorrectly transcribed EMS case numbers used to link these cases to hospital encounters, and not to any specific study or patient variable.

## DISCUSSION

Overall, the combined administration of prehospital GTN and initiation of CPAP NIV had a positive impact from both an EMS and ED/hospital perspective. The protocol led to significant improvements across multiple EMS haemodynamic parameters resulting in rapid and early symptomatic relief. While there was no evidence to support the effect of the protocol towards odds of emergency intubation, it is arguable that there are a multitude of subjective factors affecting intubation that were not measured and accounted for in this study. Of potentially greater significance was evidence to suggest a significant reduction in odds of all cause in-hospital mortality at 7 days, 30 days and in total compared with both reference groups together, and with the Regular treatment group alone. While these results should be considered in light of the study design’s limitations, they nonetheless represent important outcomes that can potentially be achieved through the early, aggressive management using simple, and inexpensive interventions.

This study is amongst the largest prehospital studies to assess the impact of a prehospital initiated intervention, assessing both prehospital and in-hospital outcomes for cardiogenic APO. The results are consistent with the literature regarding the impact on EMS vital signs and reduction in the requirement for EMS intubation ^13,16–19^. Furthermore, our study strengthens previous prehospital research which highlighted the potential impact for reduced in-hospital mortality following the initiation of prehospital treatment for acute cardiogenic APO^13,16–19^.

It is noted that Takahashi et al^20^ observed an increase in mortality for those patients with cardiogenic APO who had a prehospital time exceeding 45 minutes. This is contrast to the results of our study in which no such negative outcomes were observed despite a marginal increase in mean scene time and total mission time in the intervention group. Therefore, arguably, the negative outcomes observed by Takahashi were more likely reflective of a lack of prehospital intervention due to limitations in scope of practice than time spent within EMS.

Of additional interest to note, while the No therapy group presented seemingly less severe than the Protocol group, the initiation of prehospital treatment translated into significant mortality reduction, nonetheless. Consequently, despite their seeming heterogenous presentation, this comparison was included in the study given the potential existence of a subgroup of patients who conventionally present as “stable” who may still benefit from the initiation of early prehospital intervention.

The mandated protocol vastly outperformed the regular therapy group, both in resolution of symptom severity, and in more longer-term mortality-based outcomes. This suggests a potential synergistic effect of combining early GTN administration and CPAP initiation, given that there were a significant number of patients in the Regular therapy group that received either of these interventions, however in isolation.

## CONCLUSION

The results of this analysis provide strong evidence of the multiple benefits that can be achieved with the implementation of a treatment protocol of prehospital administered nitrates and CPAP for APO of suspected cardiac origin. In light of this, however, considerations need to be made with regards to improving and re-evaluating the accuracy of the prehospital diagnosis for cardiogenic APO.

## Data Availability

All data produced in the present study are available upon reasonable request to the authors

